## Supplementary material for "Spatial and temporal clustering of anti-SARS-CoV-2 antibodies in Illinois household cats, 2021-2023": Table 1

**Table 1.** Distribution of seropositivity of Illinois household cats across season, gender, age, and breed

|  | **Factors** | **Positive Cases (n)** | **Total (n)** | **Positive Rate (%)** |
| --- | --- | --- | --- | --- |
| **Season** | Winter 2021 (Nov-Jan) | 17 | 248 | 6.85 |
|  | Spring 2022 (Feb-April) | 58 | 539 | 10.76 |
|  | Summer 2022 (May-July) | 58 | 263 | 22.05 |
|  | Fall 2022 (Aug-Oct) | 61 | 257 | 23.74 |
|  | Winter 2022 (Nov-Jan) | 21 | 144 | 14.58 |
|  | Spring 2023 (Feb-April) | 26 | 233 | 11.16 |
| **Gender** | Male | 135 | 966 | 13.98 |
|  | Female | 106 | 734 | 14.44 |
| **Age** | Kitten (<1 year) | 8 | 27 | 29.63 |
|  | Junior (1-2 years) | 10 | 45 | 22.22 |
|  | Adult (3-6 years) | 56 | 309 | 18.12 |
|  | Senior (7-10 years) | 59 | 384 | 15.36 |
|  | Geriatric (>15 years) | 108 | 939 | 11.5 |
| **Breed** | Domestic Shorthair | 170 | 1142 | 14.89 |
|  | Domestic Longhair | 29 | 208 | 13.94 |
|  | Domestic Medium-Hair | 18 | 116 | 15.52 |
|  | Maine Coon | 5 | 31 | 16.12 |
|  | Siamese | 4 | 30 | 13.33 |
