## Supplementary material for "Spatial and temporal clustering of anti-SARS-CoV-2 antibodies in Illinois household cats, 2021-2023": Table 2

**Table 2.** Spatial and Space-Time clusters of SARS-CoV-2 infection in household cats in Illinois, United States, 2021-2023

| **Cluster Type** | **Cluster** | **Counties (n)** | **Radius (km)** | **Time Frame** | **Population** | **Observed Cases** | **Expected Cases** | **Relative Risk** | **Percent cases in cluster** | **Log-Likelihood Ratio** | **P-Value** |
| --- | --- | --- | --- | --- | --- | --- | --- | --- | --- | --- | --- |
| **Temporal** | NA | All | NA | 2022/6 - 2022/11 | 497 | 125 | 70.71 | 2.57 | 25.2 | 31.49 | 0.001 |
| **Spatial** | C1 | 3 | 45.26 | NA | 15 | 9 | 2.13 | 4.34 | 60.0 | 8.49 | 0.006 |
|  | C2 | 7 | 89.85 | NA | 28 | 12 | 3.98 | 3.12 | 42.9 | 6.89 | 0.021 |
| **Space-Time** | C1 | 8 | 78.14 | 2022/6 - 2022/9 | 28 | 18 | 3.98 | 4.80 | 64.3 | 18.87 | <0.001 |
|  | C2 | 7 | 64.22 | 2022/7 - 2022/10 | 196 | 52 | 27.89 | 2.10 | 26.5 | 11.73 | 0.013 |
|  | C3 | 5 | 46.98 | 2022/7 - 2022-10 | 6 | 6 | 0.85 | 7.18 | 100.0 | 11.76 | 0.013 |
