## Supplemental Table 1 for "Spatial and temporal clustering of anti-SARS-CoV-2 antibodies in Illinois household cats, 2021-2023"

**Table S1.** The number of seropositive and total tested samples from household cats in each county of Illinois, 2021-2023

| **County** | **Positive cases (n)** | **Total (n)** | **Positive rate (%)** |
| --- | --- | --- | --- |
| Cass | 0 | 1 | 0.00 |
| Champaign | 80 | 659 | 12.14 |
| Christian | 2 | 20 | 10.00 |
| Clark | 1 | 2 | 50.00 |
| Clinton | 0 | 2 | 0.00 |
| Coles | 6 | 40 | 15.00 |
| Cook | 29 | 190 | 15.26 |
| Cumberland | 0 | 2 | 0.00 |
| DeKalb | 1 | 1 | 100.00 |
| De Witt | 0 | 9 | 0.00 |
| Douglas | 5 | 19 | 26.32 |
| DuPage | 3 | 40 | 7.50 |
| Edgar | 3 | 12 | 25.00 |
| Effingham | 0 | 7 | 0.00 |
| Ford | 1 | 3 | 33.33 |
| Fulton | 1 | 3 | 33.33 |
| Greene | 0 | 1 | 0.00 |
| Grundy | 1 | 1 | 100.00 |
| Henry | 3 | 7 | 42.86 |
| Iroquois | 3 | 20 | 15.00 |
| Jackson | 3 | 5 | 60.00 |
| Jefferson | 0 | 1 | 0.00 |
| Kane | 1 | 5 | 20.00 |
| Kankakee | 7 | 28 | 25.00 |
| Kendall | 1 | 1 | 100.00 |
| Knox | 0 | 4 | 0.00 |
| Lake | 4 | 39 | 10.26 |
| LaSalle | 1 | 16 | 6.25 |
| Livingston | 7 | 11 | 63.64 |
| Logan | 4 | 11 | 36.36 |
| Macon | 6 | 62 | 9.68 |
| Macoupin | 2 | 5 | 40.00 |
| Madison | 1 | 17 | 5.88 |
| Marion | 0 | 2 | 0.00 |
| Marshall | 0 | 2 | 0.00 |
| McDonough | 1 | 3 | 33.33 |
| McHenry | 3 | 36 | 8.33 |
| McLean | 16 | 100 | 16.00 |
| Menard | 0 | 2 | 0.00 |
| Mercer | 0 | 1 | 0.00 |
| Montgomery | 0 | 1 | 0.00 |
| Morgan | 1 | 11 | 9.09 |
| Moultrie | 0 | 2 | 0.00 |
| Ogle | 0 | 1 | 0.00 |
| Peoria | 9 | 40 | 22.50 |
| Perry | 3 | 5 | 60.00 |
| Piatt | 1 | 26 | 3.85 |
| Richland | 1 | 5 | 20.00 |
| Rock Island | 0 | 1 | 0.00 |
| Saline | 0 | 2 | 0.00 |
| Sangamon | 12 | 74 | 16.22 |
| Shelby | 0 | 4 | 0.00 |
| St. Clair | 4 | 10 | 40.00 |
| Tazewell | 3 | 44 | 6.82 |
| Vermilion | 8 | 43 | 18.60 |
| Warren | 1 | 2 | 50.00 |
| Washington | 1 | 2 | 50.00 |
| Wayne | 0 | 5 | 0.00 |
| White | 1 | 1 | 100.00 |
| Whiteside | 0 | 1 | 0.00 |
| Will | 2 | 34 | 5.88 |
| Williamson | 1 | 3 | 33.33 |
| Woodford | 0 | 8 | 0.00 |
| **All Counties** | **244** | **1715** | **14.23** |
